# COVID-19 outbreak in Wuhan demonstrates the limitations of publicly available case numbers for epidemiological modelling

**DOI:** 10.1101/2020.04.19.20071597

**Authors:** Elba Raimúndez, Erika Dudkin, Jakob Vanhoefer, Emad Alamoudi, Simon Merkt, Lara Fuhrmann, Fan Bai, Jan Hasenauer

## Abstract

Epidemiological models are widely used to analyse the spread of diseases such as the global COVID-19 pandemic caused by SARS-CoV-2. However, all models are based on simplifying assumptions and on sparse data. This limits the reliability of parameter estimates and predictions.

In this manuscript, we demonstrate the relevance of these limitations by performing a study of the COVID-19 outbreak in Wuhan, China. We perform parameter estimation, uncertainty analysis and model selection for a range of established epidemiological models. Amongst others, we employ Markov chain Monte Carlo sampling, parameter and prediction profile calculation algorithms.

Our results show that parameter estimates and predictions obtained for several established models on the basis of reported case numbers can be subject to substantial uncertainty. More importantly, estimates were often unrealistic and the confidence / credibility intervals did not cover plausible values of critical parameters obtained using different approaches. These findings suggest, amongst others, that several models are oversimplistic and that the reported case numbers provide often insufficient information.

## Introduction

Epidemiological models are essential tools in public health as they facilitate assessments and forecasts of the spread of infectious diseases. This has been for instance demonstrated for influenza [1], dengue [2], Ebola [3], Zika [4], and – most recently – COVID-19 [5, 6]. These assessments and forecasts are the basis for political decision making [7] and therefore of vital importance.

The spectrum of mathematical modelling approaches in epidemiology ranges from relatively simple ordinary differential equation (ODE) models [8–10], partial differential equation (PDE) models [11, 12], stochastic differential equation (SDE) models [13–15], continuous-time discrete-state Markov chain (CTMC) models [15–17], to complex agent-based models [18, 19]. While ODE, PDE and SDE models provide descriptions at the population level, agent-based models are centered around formulations of the properties and dynamics of individuals. Some models explicitly account for space (usually in terms of countries, regions and/or cities) to capture spreading. Furthermore, models for the infection spread are usually combined with models of testing and reporting strategy to link them to the observed case number [20].

The choice of the modelling approach depends on the purpose of the study, the availability of information about the underlying disease and population, and the amount and quality of experimental data. Yet, all these models rely on estimates of parameter values. The parameters of epidemiological models are mostly inferred using frequentist and Bayesian parameter estimation methods. Frequentist methods often rely on parameter optimisation for obtaining point estimates and profile likelihoods for uncertainty analysis [21]. Bayesian methods exploit sampling strategies such as Markov chain Monte Carlo (MCMC) methods [20, 22] or variational inference [23]. Also flexible emulator based methods based, e.g. on Gaussian process, have been applied [24]. For applications in which competing hypotheses are available, the parameter estimation is often complemented by model selection. Established model selection measures include the Akaike Information Criterion (AIC) [25], the Bayesian Information Criterion (BIC) [26], or Bayes factors [27].

In this study, we exploit state-of-the-art parameter estimation and model selection approaches to perform an analysis of the COVID-19 outbreak in Wuhan, China. The first cases of COVID-19 were reported on December 30, 2019 and the Chinese Center for Disease Control and Prevention confirmed the isolation of a novel virus on January 7, 2020 [28]. Already by January 27, there were 1590 confirmed cases which include severe cases and 85 cumulative death cases in Wuhan, and several exported confirmed cases to Cambodia, Canada, France, Japan, Malaysia, Nepal, Republic of Korea, Singapore, Thailand, United States of America, and Vietnam [29]. As SARS-CoV-2 spread quickly, the Director-General of World Health Organisation (WHO) declared the flood of infections caused by SARS-CoV-2 a global pandemic on March 11 [30].

Our study complements other modelling efforts [31–43] by considering multiple established model types, observables, parameter estimation and model selection approaches. To recapitulate the situation in the beginning of the pandemic, we limit the use of prior knowledge to a minimum. This highlights challenges, e.g. the limited information content of case numbers and the dependence on proper model topology, but also opportunities for quantitative modelling in epidemiology.

## Results

### Observable selection and parameter identifiability

For this study, we considered the case numbers reported by the Hubei Province Health Commission and Wuhan Municipal Health Commission [44, 45]. These case numbers were particularly relevant for the analysis of the early transmission dynamics and the planning of interventions. Accordingly, these data were the basis of several modelling studies on the dynamics of COVID-19 epidemic (see e.g. [34, 38–40, 46–49]). Here, we used the time interval from January 9 to February 9, as afterwards the definition of a positive test changed [50], which limits the comparability.

The Chinese Center for Disease Control and Prevention provides time-resolved information on:

- **Reported number of infected individuals**: *y*_*I*_(*t*)
- **Reported number of recovered individuals**: *y*_*R*_(*t*)
- **Reported number of deceased individuals**: *y*_*D*_(*t*)
- **Reported cumulative number of infected individuals**: *y*_*T*_ (*t*) = *y*_*I*_(*t*) + *y*_*R*_(*t*) + *y*_*D*_(*t*)

The reported number of deceased individuals is probably most accurate, yet the overall reliability of the measurement and the distribution of the errors is unknown. As in the literature different combinations of these observables are used for model parameterisation, we consider here the following fitting scenarios:

- O1: Observations of *y*_*T*_ and *y*_*D*_
- O2: Observations of *y*_*I*_ and *y*_*D*_
- O3: Observations of *y*_*I*_, *y*_*R*_ and *y*_*D*_

As different studies considered different aspects of the data, we first asked which scenario is most suited to determine the parameters of the infection process.

To address this question, we employed a deterministic Susceptible-Exposed-Infectious-Recovered-Deceased (SEIRD) model [51]. This compartmental model describes the size of the susceptible (*S*), exposed (*E*), infectious (*I*), recovered (*R*) and deceased (*D*) subgroups (Figure 1A). The time-dependence of the subgroup sizes is governed by the ODEs:

**Figure 1:**
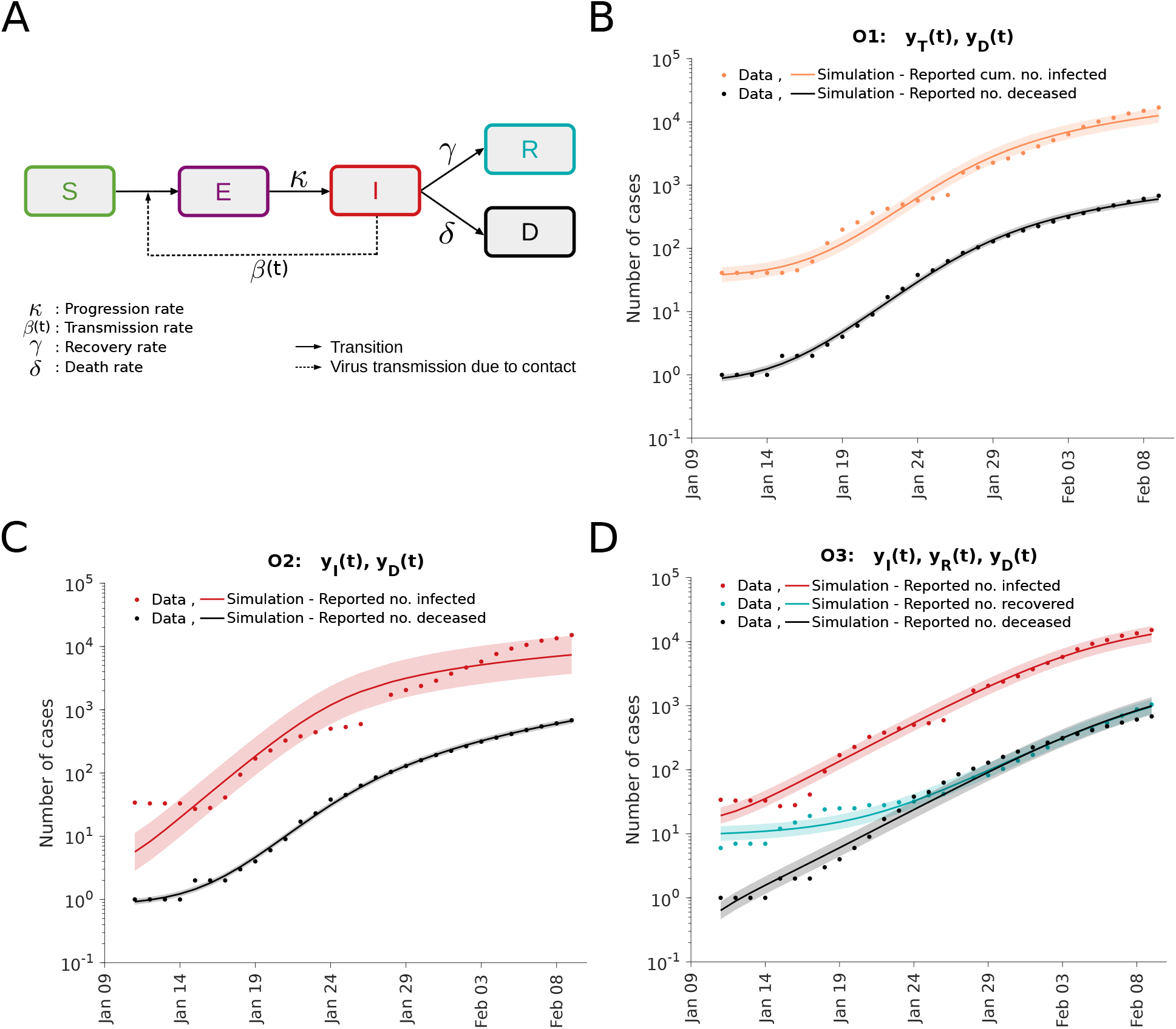
Analysis of different observable combinations. (A) Schematic of the SEIRD model. (B,C,D) Fitting results for observation scenarios O1, O2, and O3 assuming log-normally distributed measurement noise. The simulation for the maximum likelihood estimate (line) and interval for +*/−* one standard deviation of the inferred noise level (shaded area) is depicted.

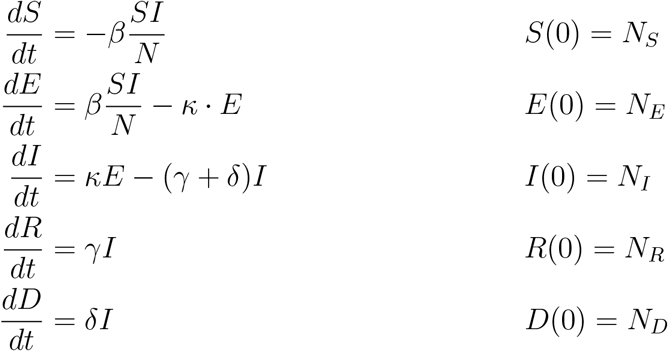

in which *β* is the average number of contacts per person per time which result in an infection, *κ* is the rate at which exposed individuals become infectious, *γ* is the rate at which infectious individuals recover, *δ* is the rate at which infectious individuals decease, and *N* = *S*(*t*) + *E*(*t*) + *I*(*t*) + *R*(*t*) + *D*(*t*) is the overall population size. Note that the inverse of the rate *κ* is the average incubation time *T*_*E*_ = *κ*^*−-1*^. The initial conditions for the different state variables are given by *N*_*S*_, *N*_*E*_, *N*_*I*_, *N*_*R*_ and *N*_*D*_. The initial conditions are usually non-zero and have to be inferred along with the unknown model parameters. Following previous work [52], we applied the simplifying assumption that infectious individuals are observed.

For all observable scenarios we performed a maximum likelihood estimation assuming normally as well as log-normally distributed measurement noise with unknown standard deviations (see *Materials and Methods*). The multi-start local optimisations converged (Supplementary Figure S1A) and the simulations with the maximum likelihood estimates achieved a good agreement with the observed data (Figure 1B-D and Supplementary Figure S1C-E). This confirms the findings of other research groups showing that the SEIRD model is sufficient to describe the dynamics of the COVID-19 outbreak in Wuhan. The comparably low noise levels inferred for the number of deceased individuals confirms our expectation that these observations are most reliable. Model selection based on AIC and BIC indicated a strong support for log-normally distributed measurement noise (Supplementary Figure S1B).

For an in-depth analysis of the impact of the choice of observables, we performed uncertainty analysis using frequentist and Bayesian methods (Figure 2). Profile likelihood calculation and MCMC sampling revealed that O3 provides improved parameter identifiability and decreased parameter uncertainties compared to O1 and O2 (Figure 2A-C). This was to be expected as O3 uses three observables (*I, R* and *D*) while O1 and O2 use two observables (and a subset of the information encoded in O3). Interestingly, the large parameter uncertainties for O1 and O2 are only partially reflected in the prediction uncertainties (Figure 2D-F) due to a strong parameter correlation (Supplementary Figure S2,S3,S4).

**Figure 2:**
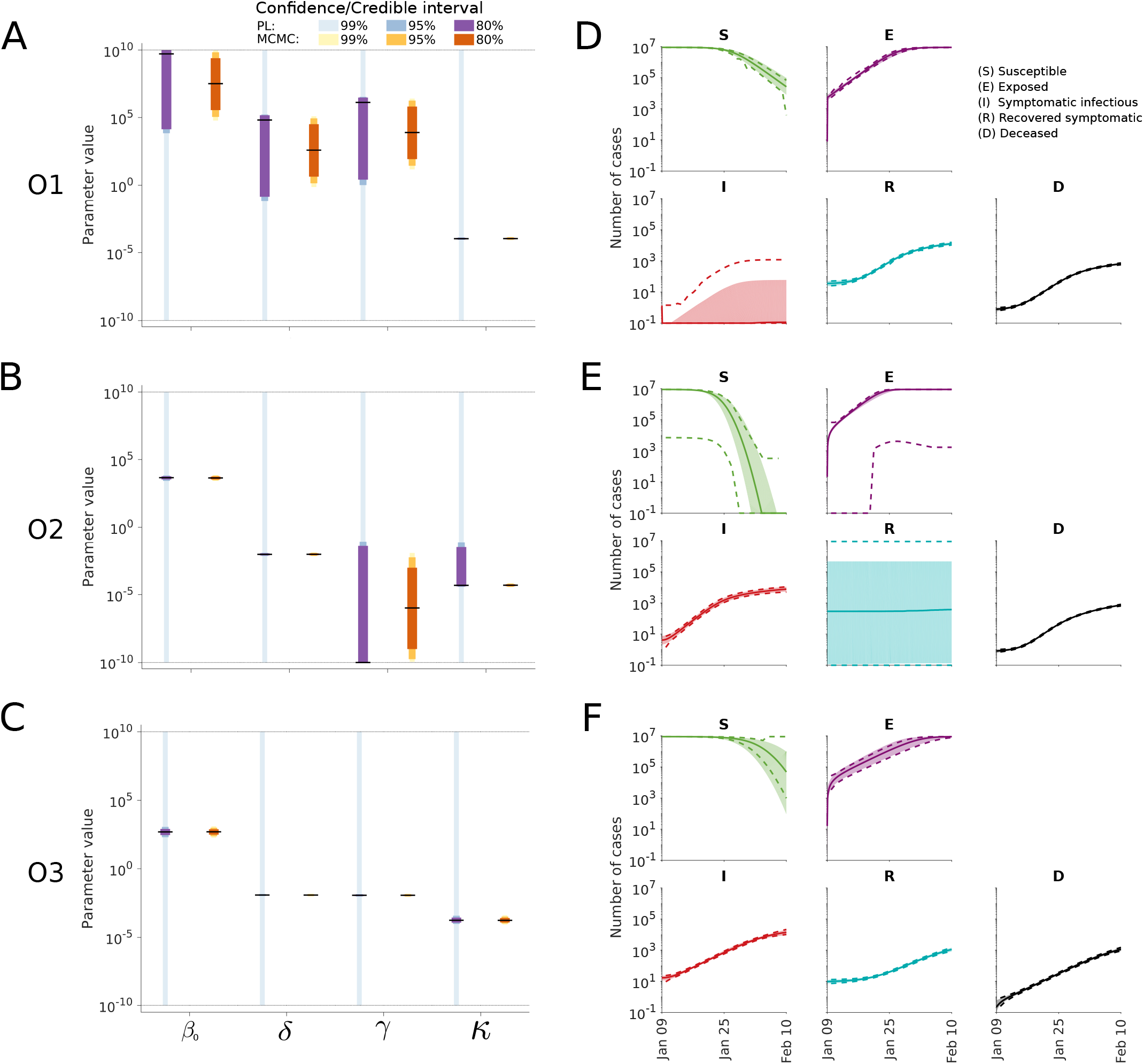
Uncertainty quantification for the different observable combinations. (A-C) Parameter confidence / credibility intervals obtained using profile calculation and MCMC samples. The gray lines indicate the employed parameter boundaries. (D-F) Posterior of state variables obtained by MCMC sampling. Medians (line) and 99% confidence (dashed lines) / credibility intervals (area) are indicated.

The most critical observation was that the parameter estimates are not realistic and that the credibility intervals derived from the MCMC samples are too narrow. The 99%-credibility intervals for O1, O2 and O3 suggested that *κ* is in the interval of [0.35, 4.11] *×* 10^−4^ days^−1^ This would imply an incubation period of [0.24, 2.83] *×* 10^4^ days. This is unrealistic and not consistent with the estimates reported by the WHO which indicate a median incubation time of 5-6 days [53]. Similar inconsistencies are observed for the basic reproduction number. Not only the Bayesian parameter estimates are off, but also the maximum likelihood estimates. However, in contrast to narrow 99%-credibility intervals computed from MCMC samples, the 99%-confidence intervals derived from profile likelihoods are broad and cover realistic values. This indicates that estimates derived from case numbers can be unrealistic and their reliability should be assessed using different approaches.

As the information encoded in the number of reported cases during the initial infection appears insufficient, we complemented it in the following with a log-normal prior for the incubation period as specified in the *Materials and Methods* section. The parameters of the prior are derived from the work of [54], which is based on the infections among travellers from Wuhan. As O3 with this prior achieves plausible estimates, we considered for the remainder this setup (Figure 3A).

**Figure 3:**
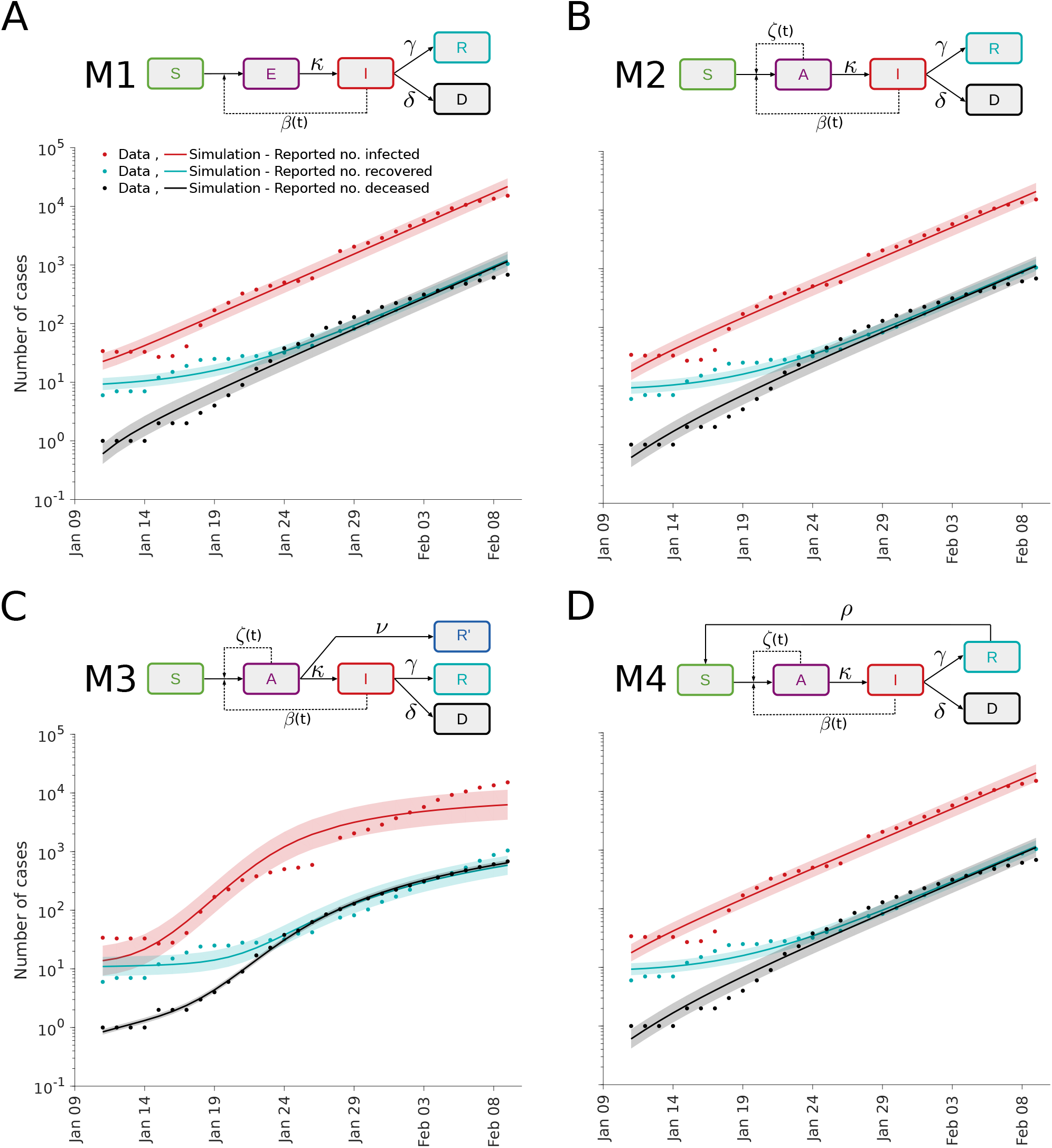
Fit for different epidemiological models. (A-D) Illustration of model structures (top) and model-data comparison (bottom). The simulation for the maximum likelihood estimate (line) and interval for +*/−* one standard deviation of the inferred noise level (shaded area) is depicted.

### Analysis of transmission process

The SEIRD model with the prior on the incubation period provides a reasonable description of the case numbers reported for Wuhan (Figure 3A). Yet, this widely used model disregards the observation that patients are asymptomatic [55]. These patients can be infectious but are more difficult to detect. To study the impact of asymptomatic patients, we consider besides the basic SEIRD model (M1) also two alternative epidemiological models:

- M2: A SAIRD model considering asymptomatic individuals (*A*) with transmission rate *ζ* and symptomatic individuals (*I*) with transmission rate *β*. The asymptomatic individuals are assumed to become symptomatic.
- M3: A SAIRD model similar to M2, which allows for the direct recovery of asymptomatic patients. The recovered asymptomatic patients (*R*^′^) are assumed to remain unreported.

We assume that the reported number of infected individuals corresponds to the number of symptomatic patients (*I*). Accordingly, the reported number of recovered individuals is assumed to count only previously symptomatic individuals.

As a further model extension, we consider the possibility of waning immunity. A recent study suggested that the infection with SARS-CoV-2 does not necessarily induce a long-lived antibody response [56]. This has, to the best of our knowledge, not yet been evaluated using model-based approaches. To address this, we consider:

- M4: A SAIRD model similar to M2, which allows recovered individuals (*R*) to become susceptible (*S*) with rate *ρ*.

The parameters of models M1-M4 were estimated using multi-start local optimisation. The simulations of models M1 to M4 for the respective maximum a posterior estimate show a reasonable agreement with the data (Figure 3). Interestingly, while the simulations for M1, M2 and M4 are similar, the simulation for M3 shows an early saturation (Figure 3C). The reason is that the initial number of unobserved recovered patients (which can also be interpreted as immune patients) is estimated to be very high, which does not appear to be plausible.

As the fitting results for all models were highly reproducible (Figure 4A), we evaluated the AIC and BIC for model selection (Figure 4B,C). As all models achieved a relatively similar fit, the complexity penalisation has a critical impact and the ranking differs for AIC and BIC, suggesting that the differences are minor. Only M4 was consistently not supported by the available data. The model selection revealed again the limited information content of the case numbers as there is clear evidence for the relevance of asymptomatic cases.

**Figure 4:**
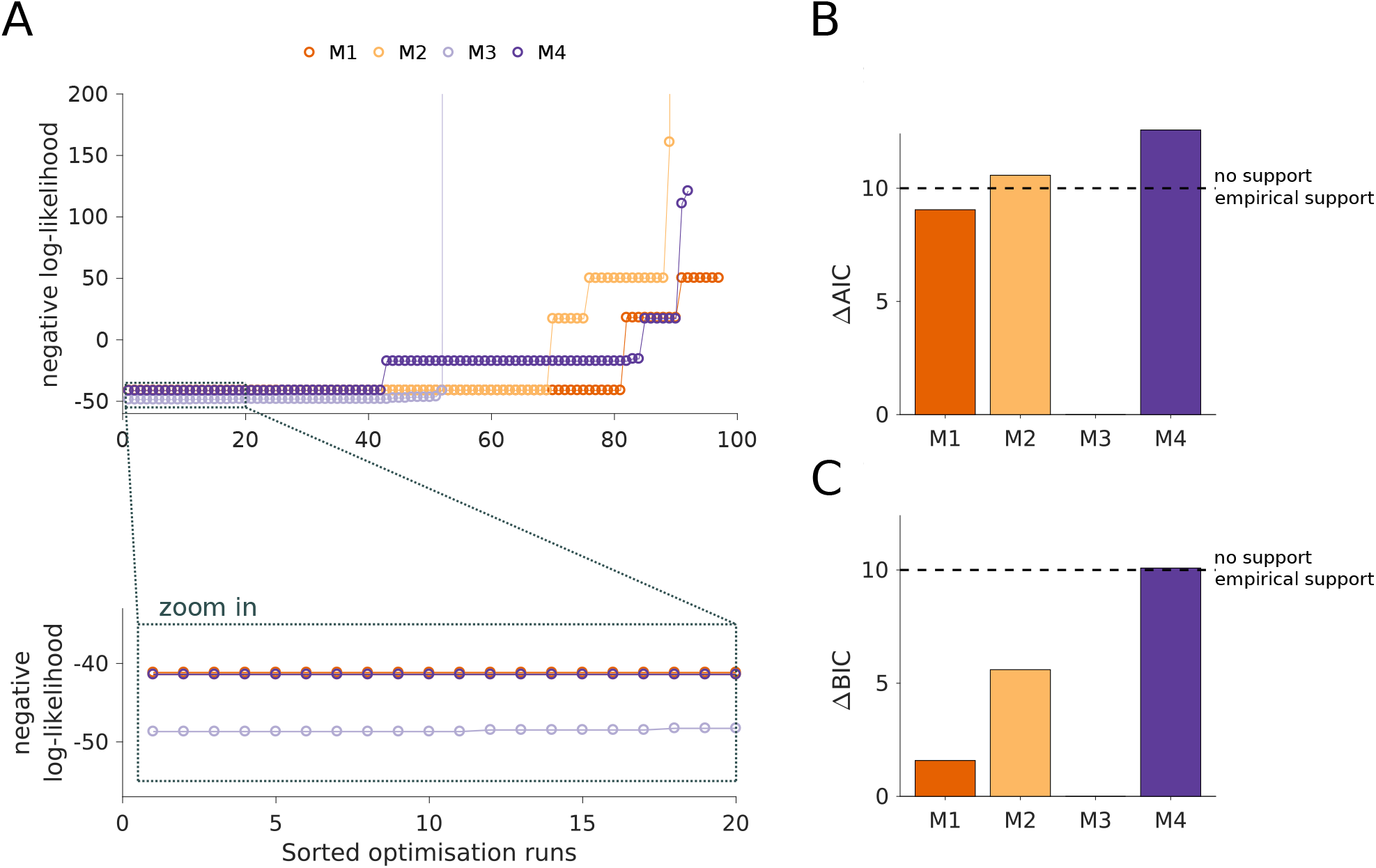
Analysis of model structure. (A) Waterfall plots for the best 100 out of 200 multi-start runs. The optimisation runs are sorted by increasing negative log-likelihood value. The lower panel shows a magnification of the best 20 starts. (B) Differences in AIC with respect to the lowest value indicate M3 as the most plausible model. (C) Differences in BIC with respect to the lowest value indicate M3 as the most plausible model. Black dashed line in (B) and (C) depicts a change of 10 units considered as a rejection threshold.

To assess the uncertainty of the parameter estimates and predictions, we computed the profile likelihoods and performed MCMC sampling (Figure 5A-D). The results indicate that the parameter uncertainties for M2-M4 are larger than for M1, but that most parameters are well determined. The profile likelihoods yield overall more conservative estimates than the sampling. The predictions of the state variables based on the sampling suggest low uncertainties of all model states (Figure 5E-H). In particular for M3 this appears unrealistic as there are so far no reports about a large number of immune individuals (Figure 5G).

**Figure 5:**
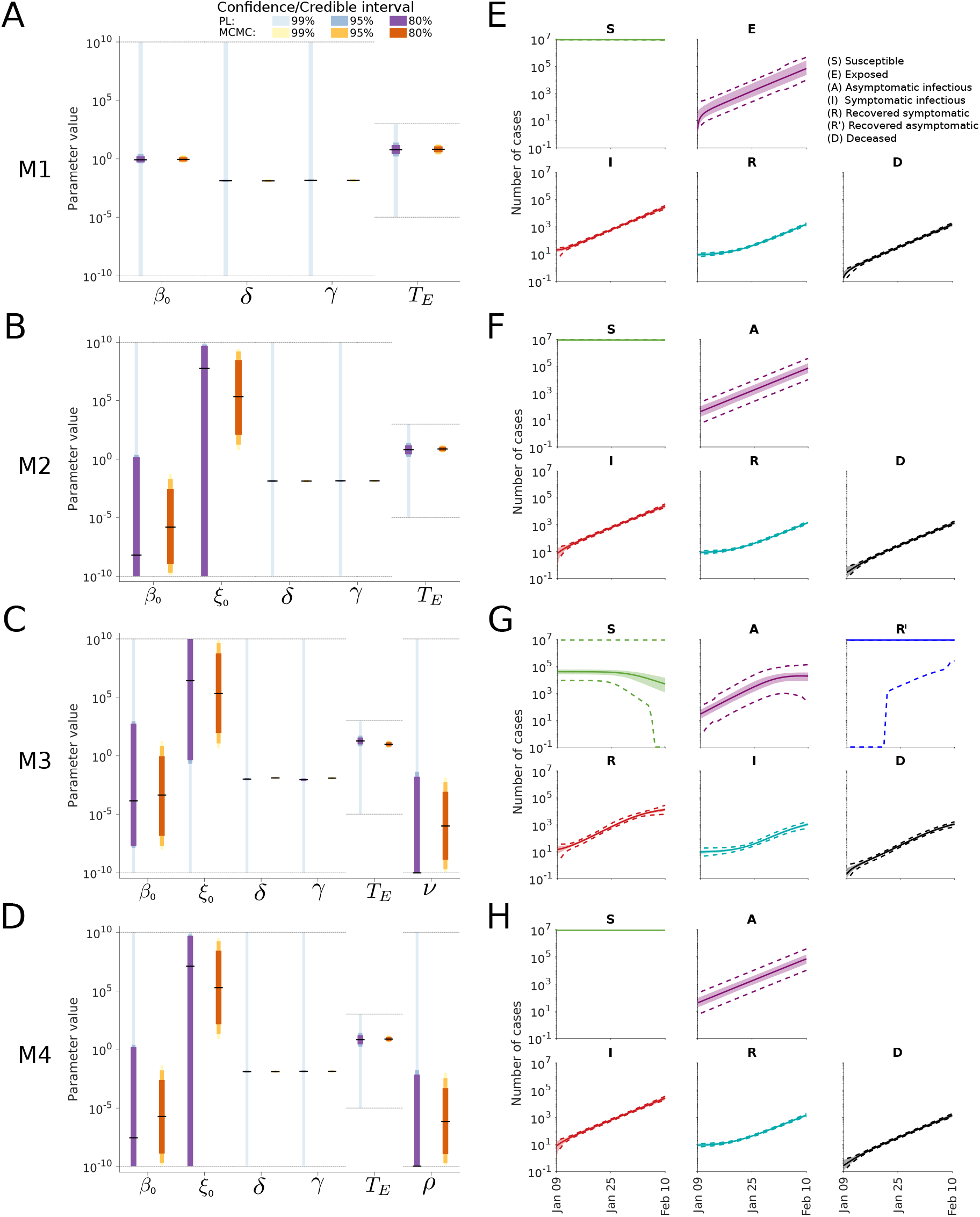
Uncertainty quantification for different model structures. (A-D) Confidence / credibility intervals for the model parameters obtained using profile calculation and MCMC sampling. The gray lines indicate the employed parameter boundaries. (E-H) Confidence / credibility intervals for the state variables obtained using prediction profile likelihood calculation and MCMC sampling. Medians (line) and 99% confidence (dashed lines) / credibility intervals (area) are indicated.

### Analysis of intervention effect

A key question in many recent studies is how much interventions such as compulsory masks and social contact restrictions, as well as the rising public awareness impact the transmission rate of SARS-CoV-2. To study how well this question can be assessed based on early case report data, we considered three simple scenarios:

- **No change** of the transmission rate.
- **Discrete change** in the transmission rate due to the compulsory masks introduced by the government in Wuhan on January 22 and substantially increasing contract restrictions.
- **Continuous change** in the transmission rate due to rising public awareness and a broad spectrum of interventions.

These scenarios are illustrated in Figure 6 and a detailed mathematical description is provided in the *Materials and Methods* section.

**Figure 6:**
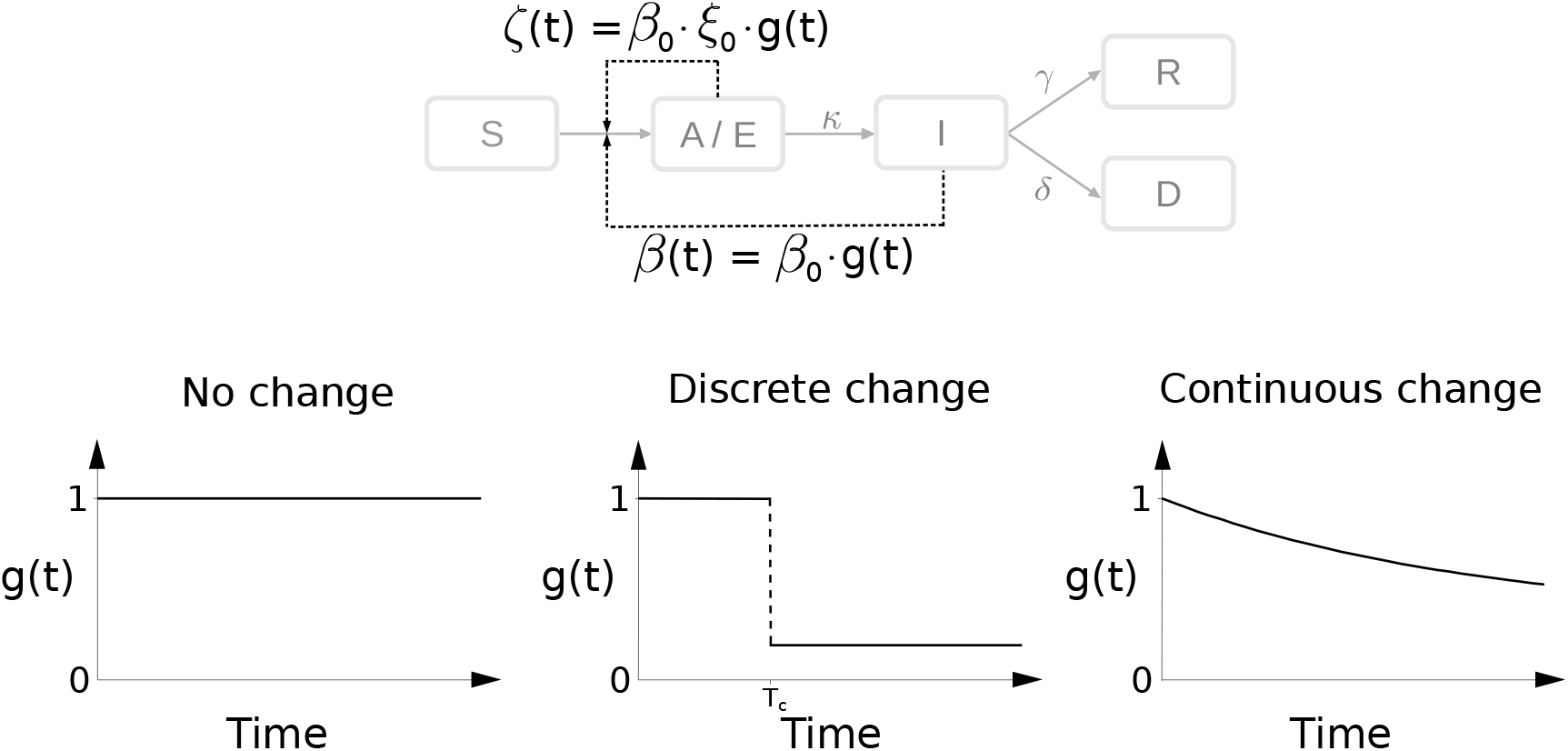
Modeling intervention strategies. Schematic of distinct intervention effects influencing *β* and *ζ. T*_*C*_ denotes the time at which an intervention is performed.

As the result of the analysis of the infection dynamics carried out in the previous section was inconclusive, we considered model M1 to M4. For all 12 combinations of model structures and intervention effects, we performed parameter estimation and uncertainty analysis. The assessment of the model selection criteria provided support for a discrete change in the transmission rate on January 22 (Figure 7A). The resulting model provides an accurate description of the data and suggests that the transmission rate dropped by around 71% (Figure 7B). The uncertainty estimates for the decrease (Δ) depend heavily on the analysis approach. While MCMC sampling yields a 99% credibility interval from 37.9 to 76.4%, the profiles suggests a much broader regime (Figure 7C). Accordingly, the reported case number for the early outbreak were not sufficient for an accurate assessment of all model parameters. Despite the parameter uncertainties, the states seem to be relatively well determined (Figure 7D).

**Figure 7:**
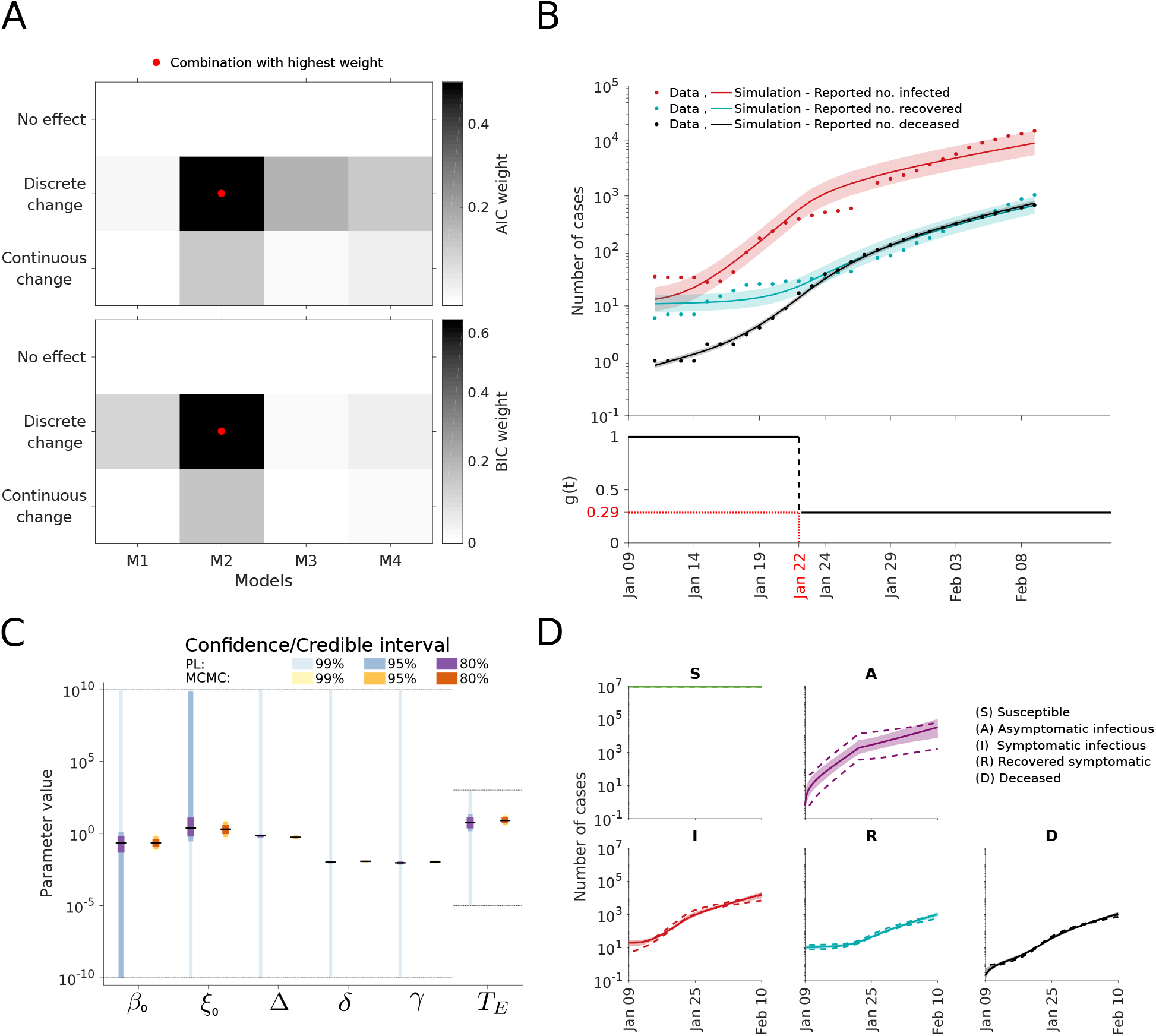
Analysis of intervention effects. (A) Model selection using AIC weights (top) and BIC weights (bottom). (B) Best model fit (top) and estimated intervention effect (bot-tom). The simulation for the maximum likelihood estimate (line) and interval for +*/−* one standard deviation of the inferred noise level (shaded area) is depicted. (C) Confidence / credibility intervals for the model parameters obtained using profile calculation and MCMC sampling. The gray lines indicate the employed parameter boundaries. (D) Confidence / credibility intervals for the state variables obtained using prediction profile likelihood calculation and MCMC sampling. Medians (line) and 99% confidence (dashed line) / credibility intervals (area) are indicated.

## Discussion

Pandemics pose a global challenge and show the importance of model-based forecasting. Forecasts influence the political decision-making process and have a significant impact on our society. Minimising model uncertainties and properly evaluating them is therefore crucial. Yet, many publications are still only using reported case number and/or omit an identifiability and uncertainty analysis [33–35, 39, 57–60]. Here, we demonstrated that both aspects are problematic.

Our analysis of the COVID-19 outbreak in Wuhan demonstrates that the parameterisation of epidemiological models can result in incorrect parameter estimates and predictions. Surprisingly, even Bayesian uncertainty analysis using MCMC sampling resulted in an underestimation of the indeterminacy and provided inaccurate predictions. In our opinion there are two reasons for this:

1. The models considered here and used in various other publications are too simple. They neglect for instance the stochastic nature of the process [61, 62], the heterogeneity of the population (e.g. the age structure [63]), time-dependent testing and reporting protocols [50], and particularities of the process (e.g. a large number of asymptomatic cases [64]). As the parameter estimates depend on the model characteristics, oversimplifications can result in biased estimates and predictions.
2. The case report data provide only limited information about the process, in particular the distribution of inter-event times are difficult of reconstruct.

Besides parameter estimation, the aforementioned limitations of case numbers were observed in the model selection process. The data did, for instance, not allow to unravel that a large number of the asymptomatic cases is not detected.

We observed that detailed prior information is required if merely case numbers are employed for parameter estimation. While literature-based priors are used in many manuscripts [65], we hypothesise that it would be better to use information about individual cases for parameter estimation and model selection. In particular the date of the onset of symptoms, the date of the positive test, and the date of recovery/death for individuals is highly relevant. These data are being collected and analysed [54, 66–68], but should in the future be shared much earlier. Furthermore, randomised testing would be required, ideally using antibody tests to determine the fraction of completely asymptomatic patients. Such studies are usually not possible during an initial phase of a pandemic but are now on the way [69].

This study does not offer new insights into the COVID-19 pandemic. However, it pin-points important pitfalls and showcases the relevance of the underlying assumptions and the available data. Furthermore, it demonstrates that even a proper uncertainty analysis using state-of-the-art frequentist or Bayesian approaches does not ensure that the true parameters and dynamics are captured within the uncertainty bounds. While we demonstrate this aspect here for deterministic compartmental models – which are the basis of many modelling studies for COVID-19 and beyond – it certainly holds also for other modelling approaches. Similarly, model-free studies are based on assumptions, data and statistical models, rendering them subject to at least the same limitations.

## Materials and Methods

### Data

The study is based on official reports on the total number of cases and the numbers of infected individuals, recovered individuals and deceased individuals. From January 11 to 20, the reports were made available by the Wuhan Municipal Health Commission [45]. Afterwards, the reports were organised by the Health Commission of Hubei Province [44]. The complete data sets are listed in Table 1.

**Table 1:**
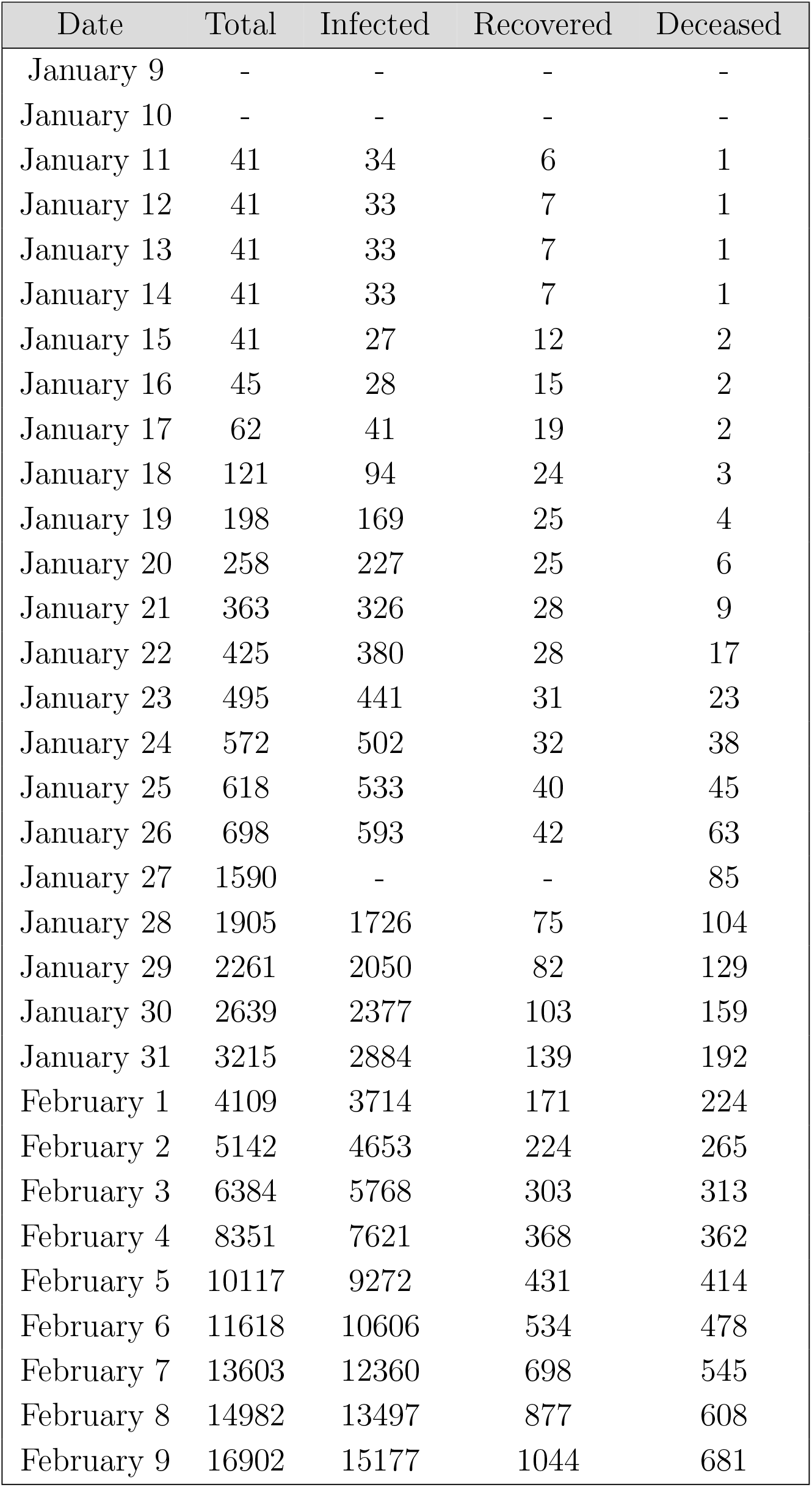
Number of total, infected, deceased and recovered cases in Wuhan, China. Missing data in the official reports are indicated by “-”.

The exact population size in the city of Wuhan in the period under consideration is not precisely known due to Chinese New Year. In this study we assume a population size of 9 million which was mentioned by the mayor of Wuhan, Zhou Xianwang [70].

### Mathematical models

We considered four different deterministic compartmental models for the description of the transmission process. The state variables of the models are the number of individuals with particular characteristics and the notations can be found in Table 2.

**Table 2:**
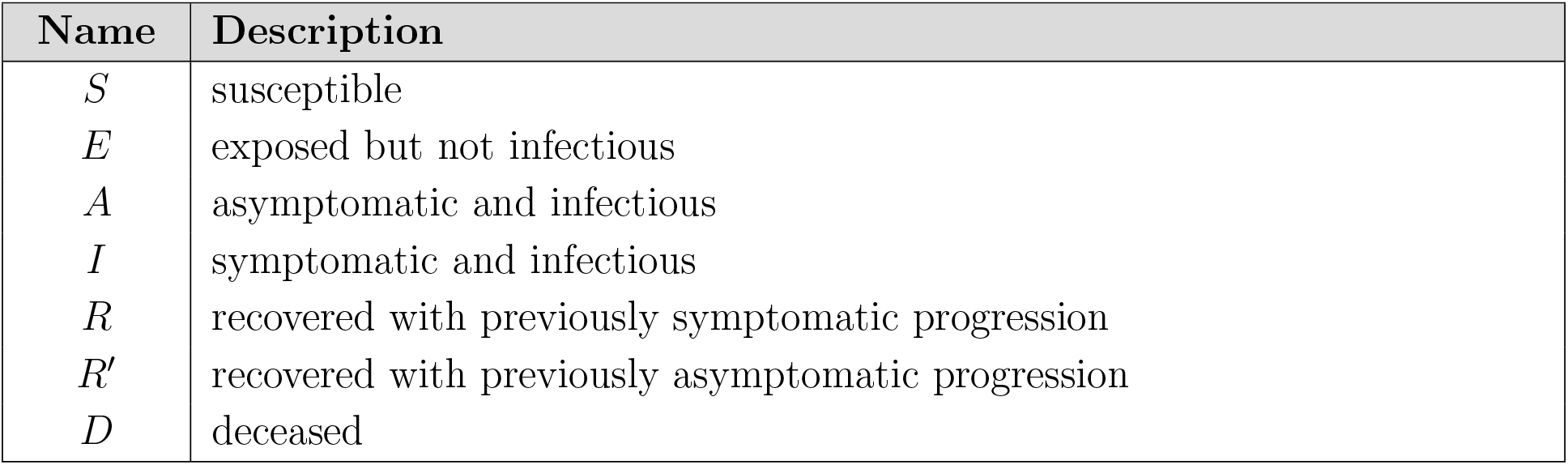
State variables of the mathematical models.

| Name | Description |
| --- | --- |
| $S$ | susceptible |
| $E$ | exposed but not infectious |
| $A$ | asymptomatic and infectious |
| $I$ | symptomatic and infectious |
| $R$ | recovered with previously symptomatic progression |
| $R'$ | recovered with previously asymptomatic progression |
| $D$ | deceased |

The models allow for various processes which result in the transitions of individuals between compartments (see Figure 3). A description of the rates is provided in Table 3. For the time-dependence of the transmission rates *β* And *ζ*,

**Table 3:** Rates in the mathematical models.

| Name | Process | Description |
| --- | --- | --- |
| $\beta(t)$ | $S + I \rightarrow E/A$ | (time-dependent) transmission rate for symptomatic |
| $\zeta(t)$ | $S + A \rightarrow E/A$ | (time-dependent) transmission rate for asymptomatic |
| $\kappa$ | $E/A \rightarrow I$ | progression rate ($= T_E^{-1}$ for incubation time $T_E$) |
| $\gamma$ | $I \rightarrow R$ | recovery rate for symptomatic case |
| $\nu$ | $A \rightarrow R'$ | recovery rate for asymptomatic case |
| $\delta$ | $I \rightarrow D$ | death rate |
| $\rho$ | $R \rightarrow S$ | rate at which immunity wanes |

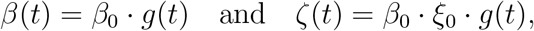

we considered three scenarios:

- No change: *g*(*t*) = 1
- Discrete change: 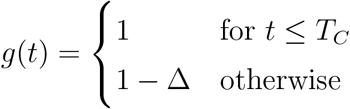
- Continuous change: *g*(*t*) = *e*^*−kt*^

The function *g*(*t*) describes the reduction of the transmission rates compared to baseline at *t* = 0, with *g*(0) = 1 for all scenarios. The parameters for the discrete change are the time point *T*_*C*_ and the relative reduction Δ, and for the continuous change we have the decay rate *k*. The parameter 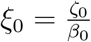 denotes the relative difference between the transmission rates of the symptomatic individuals (*β*(*t*)) and asymptomatic individuals (*ζ*(*t*)).

The ODEs governing the dynamics of the different compartmental models are provided in Table 5. We decided to initialise the model on January 9 since which the virus was detected [71], defining *t* = 0. As on January 22 the wearing of masks became mandatory, we set for the scenario of a discrete reduction of *β* for *T*_*C*_ = 13 days.

**Table 4:**
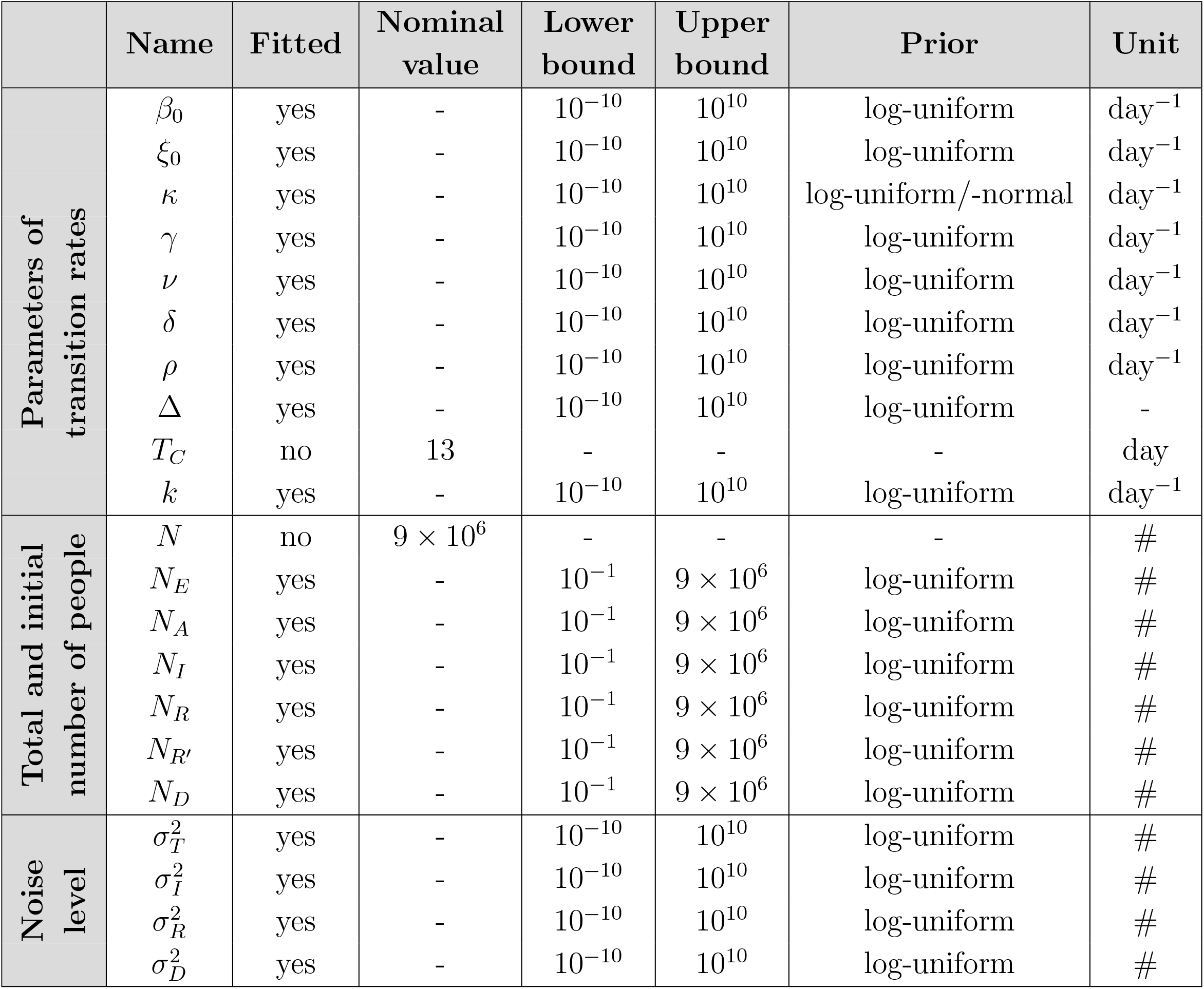
Model parameters. Nominal values, lower bounds, upper bounds, priors and units for the model parameters. The nominal values are only used for model parameters which are not fitted.

|  | Name | Fitted | Nominal value | Lower bound | Upper bound | Prior | Unit |
| --- | --- | --- | --- | --- | --- | --- | --- |
| Parameters of transition rates | $\beta_0$ | yes | - | $10^{-10}$ | $10^{10}$ | log-uniform | day<sup>-1</sup> |
| | $\xi_0$ | yes | - | $10^{-10}$ | $10^{10}$ | log-uniform | day<sup>-1</sup> |
| | $\kappa$ | yes | - | $10^{-10}$ | $10^{10}$ | log-uniform/-normal | day<sup>-1</sup> |
| | $\gamma$ | yes | - | $10^{-10}$ | $10^{10}$ | log-uniform | day<sup>-1</sup> |
| | $\nu$ | yes | - | $10^{-10}$ | $10^{10}$ | log-uniform | day<sup>-1</sup> |
| | $\delta$ | yes | - | $10^{-10}$ | $10^{10}$ | log-uniform | day<sup>-1</sup> |
| | $\rho$ | yes | - | $10^{-10}$ | $10^{10}$ | log-uniform | day<sup>-1</sup> |
| | $\Delta$ | yes | - | $10^{-10}$ | $10^{10}$ | log-uniform | - |
| | $T_C$ | no | 13 | - | - | - | day |
| | $k$ | yes | - | $10^{-10}$ | $10^{10}$ | log-uniform | day<sup>-1</sup> |
| Total and initial number of people | $N$ | no | $9 \times 10^6$ | - | - | - | # |
| | $N_E$ | yes | - | $10^{-1}$ | $9 \times 10^6$ | log-uniform | # |
| | $N_A$ | yes | - | $10^{-1}$ | $9 \times 10^6$ | log-uniform | # |
| | $N_I$ | yes | - | $10^{-1}$ | $9 \times 10^6$ | log-uniform | # |
| | $N_R$ | yes | - | $10^{-1}$ | $9 \times 10^6$ | log-uniform | # |
| | $N_{R'}$ | yes | - | $10^{-1}$ | $9 \times 10^6$ | log-uniform | # |
| | $N_D$ | yes | - | $10^{-1}$ | $9 \times 10^6$ | log-uniform | # |
| Noise level | $\sigma_T^2$ | yes | - | $10^{-10}$ | $10^{10}$ | log-uniform | # |
| | $\sigma_I^2$ | yes | - | $10^{-10}$ | $10^{10}$ | log-uniform | # |
| | $\sigma_R^2$ | yes | - | $10^{-10}$ | $10^{10}$ | log-uniform | # |
| | $\sigma_D^2$ | yes | - | $10^{-10}$ | $10^{10}$ | log-uniform | # |

**Table 5:** Mathematical models. ODEs for the transmission dynamics, initial conditions and observations functions are defined for all considered model structures. As some models consider only a subset of the state variables, some rows are empty.

|  | SEIRD model (M1) | SAIRD model (M2) | SAIRRD model (M3) | SAIRDS model (M4) |
| --- | --- | --- | --- | --- |
| Infection dynamics | $\frac{dS}{dt} = -\beta \frac{SI}{N}$ $\frac{dE}{dt} = \beta \frac{SI}{N} - \kappa E$ $\frac{dI}{dt} = \kappa E - (\gamma + \delta)I$ $\frac{dR}{dt} = \gamma I$ $\frac{dD}{dt} = \delta I$ | $\frac{dS}{dt} = -\beta \frac{SI}{N} - \zeta \frac{SA}{N}$ $\frac{dA}{dt} = \beta \frac{SI}{N} + \zeta \frac{SA}{N} - \kappa A$ $\frac{dI}{dt} = \kappa A - (\gamma + \delta)I$ $\frac{dR}{dt} = \gamma I$ $\frac{dD}{dt} = \delta I$ | $\frac{dS}{dt} = -\beta \frac{SI}{N} - \zeta \frac{SA}{N}$ $\frac{dA}{dt} = \beta \frac{SI}{N} + \zeta \frac{SA}{N} - (\kappa + \nu)A$ $\frac{dI}{dt} = \kappa A - (\gamma + \delta)I$ $\frac{dR}{dt} = \gamma I$ $\frac{dR'}{dt} = \nu A$ $\frac{dD}{dt} = \delta I$ | $\frac{dS}{dt} = -\beta \frac{SI}{N} - \zeta \frac{SA}{N} + \rho R$ $\frac{dA}{dt} = \beta \frac{SI}{N} + \zeta \frac{SA}{N} - \kappa A$ $\frac{dI}{dt} = \kappa A - (\gamma + \delta)I$ $\frac{dR}{dt} = \gamma I - \rho R$ $\frac{dD}{dt} = \delta I$ |
| Initial conditions | $S(0) = N - N_E - N_I - N_R - N_D$ $E(0) = N_E$ $I(0) = N_I$ $R(0) = N_R$ $D(0) = N_D$ | $S(0) = N - N_A - N_I - N_R - N_D$ $A(0) = N_A$ $I(0) = N_I$ $R(0) = N_R$ $D(0) = N_D$ | $S(0) = N - N_A - N_I - N_R - N_{R'} - N_D$ $A(0) = N_A$ $I(0) = N_I$ $R(0) = N_R$ $R'(0) = N_{R'}$ $D(0) = N_D$ | $S(0) = N - N_A - N_I - N_R - N_D$ $A(0) = N_A$ $I(0) = N_I$ $R(0) = N_R$ $D(0) = N_D$ |
| Observables | $y_T = I + R + D$ $y_I = I$ $y_R = R$ $y_D = D$ | $y_T = I + R + D$ $y_I = I$ $y_R = R$ $y_D = D$ | $y_T = I + R + D$ $y_I = I$ $y_R = R$ $y_D = D$ | $y_T = I + R + D$ $y_I = I$ $y_R = R$ $y_D = D$ |

The state variables of the model were linked to the observed case numbers using observation functions. The observation functions for the total number of cases (*y*_*T*_) as well as for the number of infected (*y*_*I*_), recovered (*y*_*R*_) and deceased (*y*_*D*_) people are provided in Table 5. As the reported numbers are subject to unknown measurement noise, we considered two error models:

- Additive normally distributed measurement noise:

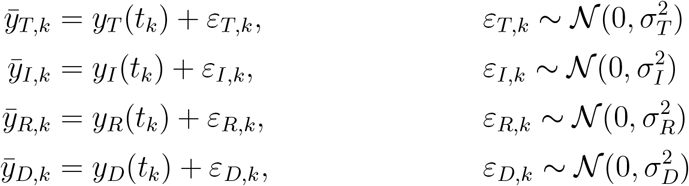
- Multiplicative log-normally distributed measurement noise:

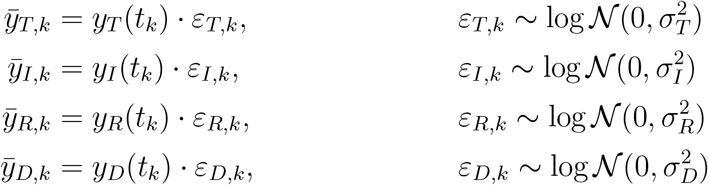

The observation time points *t*_*k*_, *k* = 1, …, 32, are the days listed in Table 1, and the measurements (as indicated with the superscript *m*) are the respective case numbers. The distribution parameters *σ*_*T*_, *σ*_*I*_, *σ*_*R*_ and *σ*_*D*_ were considered as unknown.

In the following the parameters of the transition rates, the total and initial number of people in different compartments, and the parameters of the noise distribution are inferred. A comprehensive list of all model parameters and implemented constraints is provided in Table 4. The boundaries of the search space were chosen very conservatively to indicate the initial knowledge gap about SARS-CoV-2 and COVID-19.

### Parameter estimation

To infer the unknown model parameters we used frequentist and Bayesian approaches. These approaches considered the conditional probability *p*(*𝒟*|*θ*) of the data *𝒟* given the parameters *θ*, also known as likelihood. For additive normally distributed measurement noise the likelihood function is

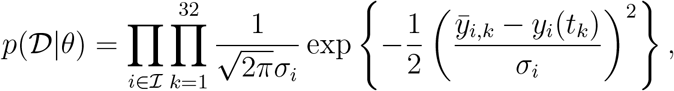

and for multiplicative log-normally distributed measurement it is

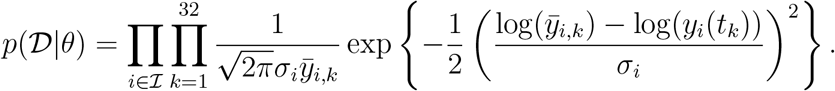

The set of considered observables is encoded by *ℐ* and differs for the scenarios O1 to O3: *ℐ*_O1_ = *{T, D}, ℐ*_O2_ = *{I, D}*, and *ℐ*_O3_ = *{I, R, D}*. In addition to the case reports, we also incorporated knowledge available before the parameter estimation. For Bayesian approaches this was done by defining a prior distributions. These prior distributions are mostly log-uniform with conservative upper and lower bounds, meaning that the distribution over the log-transformed parameter values is flat. For *κ* we include in parts of the study information about the incubation period [54], given by a log-normal prior:

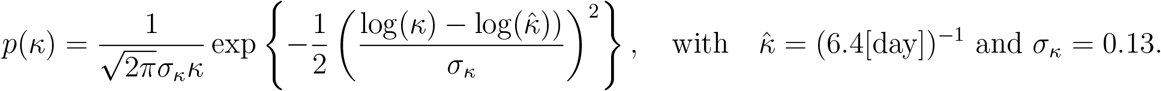

For the frequentist approaches the available estimate of *κ* is treated as a data point.

*Remark:* For the parameter estimation we consider the log-transformed parameter values. For the log-transformed parameters, the log-uniform prior become effectively a uniform prior. This renders the frequentist and the Bayesian approaches comparable, namely, the maximum likelihood and the maximum a posteriori estimates coincide.

#### Maximum likelihood and maximum a posteriori estimates

To determine the maximum likelihood and the maximum a posteriori estimates, we minimised the negative log-likelihood function and negative log-posterior function, respectively. As these optimisation problems are non-linear and non-convex, we used multi-start local optimisation. The starting points for the local optimisations were generated using latin hypercube sampling. Local optimisation was performed using the interior point algorithm implemented in the MATLAB function `lsqnonlin.m`, which exploits the least-squares like structure of the optimisation problems. To facilitate convergence, we computed the gradients of the residuals via forward sensitivity equations. The convergence of the global optimisation was assessed using waterfall plots.

For each of the 18 considered combinations of compartment model, noise model, observable scenario and intervention scenario, we performed 200 local optimisations. For all combinations at least 15 runs converged to the observed global optimum. This suggested that the results are highly reliable.

#### Frequentist uncertainty analysis

To evaluate the (frequentist) parameter and prediction confidence intervals we used profile likelihoods [72, 73]. The profile likelihoods were computed using the MATLAB toolbox Data2Dynamics [74]. This toolbox implements optimisation-based methods with adaptive step-size selection as well as fast integration-based methods [75]. To ensure the robustness of the results, the consistency of the outcomes was checked and confirmed.

The profile likelihoods were used to derive the (finite sample) confidence intervals. For a significance level *α*, the bounds of the confidence interval were determined as the smallest and largest parameter values for which the profile likelihood stays above the threshold defined by the *α*th-percentile of the *χ*^2^-distribution [76]. We used the *χ*^2^-distribution with one degree of freedom which yields the so called point-wise confidence intervals.

#### Bayesian uncertainty analysis

To evaluate the (Bayesian) parameter and prediction credibility intervals we used Markov chain Monte Carlo sampling [77]. The parameter posterior distribution was sampled using the Adaptive Metropolis algorithm implemented in the MATLAB toolbox PESTO [78]. The methods are self-tuning and provided good convergence properties. Convergence after burn-in was assessed using the Geweke test [79]. The parameter samples were used to generate samples for the model states and observables.

The samples of parameters and predictions were used to derive the credibility intervals. For a credibility level *α*, the bounds of the credibility interval were determined as the 100*α/*2-and the 100(1 − *α/*2)-percentile of the respective sample. This procedure yields the so called equal-tailed interval.

### Model selection

We considered competing hypotheses on the dynamics of the infection process, the effect of the intervention and the noise distribution. Each of the resulting models was assessed using the Akaike information criterion (AIC),

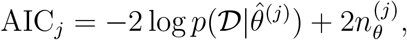

and the Bayesian information criterion (BIC),

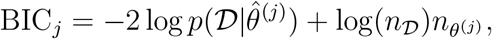

in which *j* is the model index, 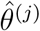 is the maximum likelihood estimate for the *j*-th model, and *n*_*θ*_(*j*) is the number of parameter of the *j*-th model. The number of independent data points is denoted by *n*_*D*_. AIC and BIC account for the likelihood of the data and penalise model complexity. Low AIC and BIC values are favorable. We consider a difference 10 between AIC/BIC values of different models as substantial [80].

For analysis and visualisation we also evaluated the AIC and BIC weights [80]. The AIC weight for the *j*-th model is defined as

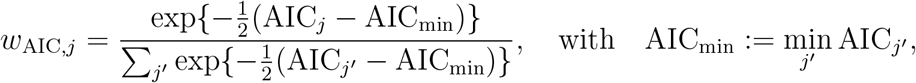

and provides the weight of evidence in favor of the *j*-th model being the actual best model in terms of the Kullback–Leibler Information (assuming that the true model is in the considered set). The BIC weight for the *j*-th model is defined as

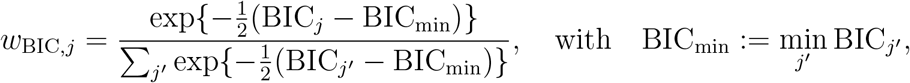

and provides an approximation to the Bayesian posterior probability of the *j*-th model. AIC and BIC weights are between 0 and 1, and a high value indicates a strong support.

### Implementation and availability

The model formulation, parameter estimation and profile likelihoods were performed in the MATLAB toolbox Data2Dynamics (https://github.com/Data2Dynamics/d2d) [74]. Outliers in the computed prediction profiles arising from calculation error were corrected subsequently. The calculation of parameter confidence intervals and MCMC sampling was carried out using the MATLAB toolboxes PESTO (https://github.com/ICB-DCM/PESTO) [78] and AMICI (https://github.com/ICB-DCM/AMICI) [81, 82]. For numerical integration Data2Dynamics and AMICI rely on the SUNDIALS solver CVODES [83].

The complete implementation (including the respective version of the used toolboxes) and data are available on ZENODO (https://doi.org/10.5281/zenodo.3757227). This includes the MATLAB code as well as the the specification of the parameter estimation problems as PEtab files [84] (with the model in SBML format [85]).

## Data Availability

The complete implementation (including the respective version of the used toolboxes) and data are available on ZENODO (https://doi.org/10.5281/zenodo.3757227). This includes the MATLAB code as well as the the specification of the parameter estimation problems as PEtab files (with the model in SBML format).

## Author contributions

J.H., F.B. designed the study. E.R., E.D., J.V. implemented the models. E.R., E.D. implemented the pipeline. F.B. extracted and formatted data. E.R., E.D., performed parameter estimation and model selection. E.A., L.F. researched the literature. S.M. implemented the models in the PEtab format. E.R., E.D., J.H. drafted the manuscript. All authors discussed and approved the study, and revised and approved the final manuscript.

## Additional Information

No competing interests.

## Supplementary Figures

**Figure S1:**
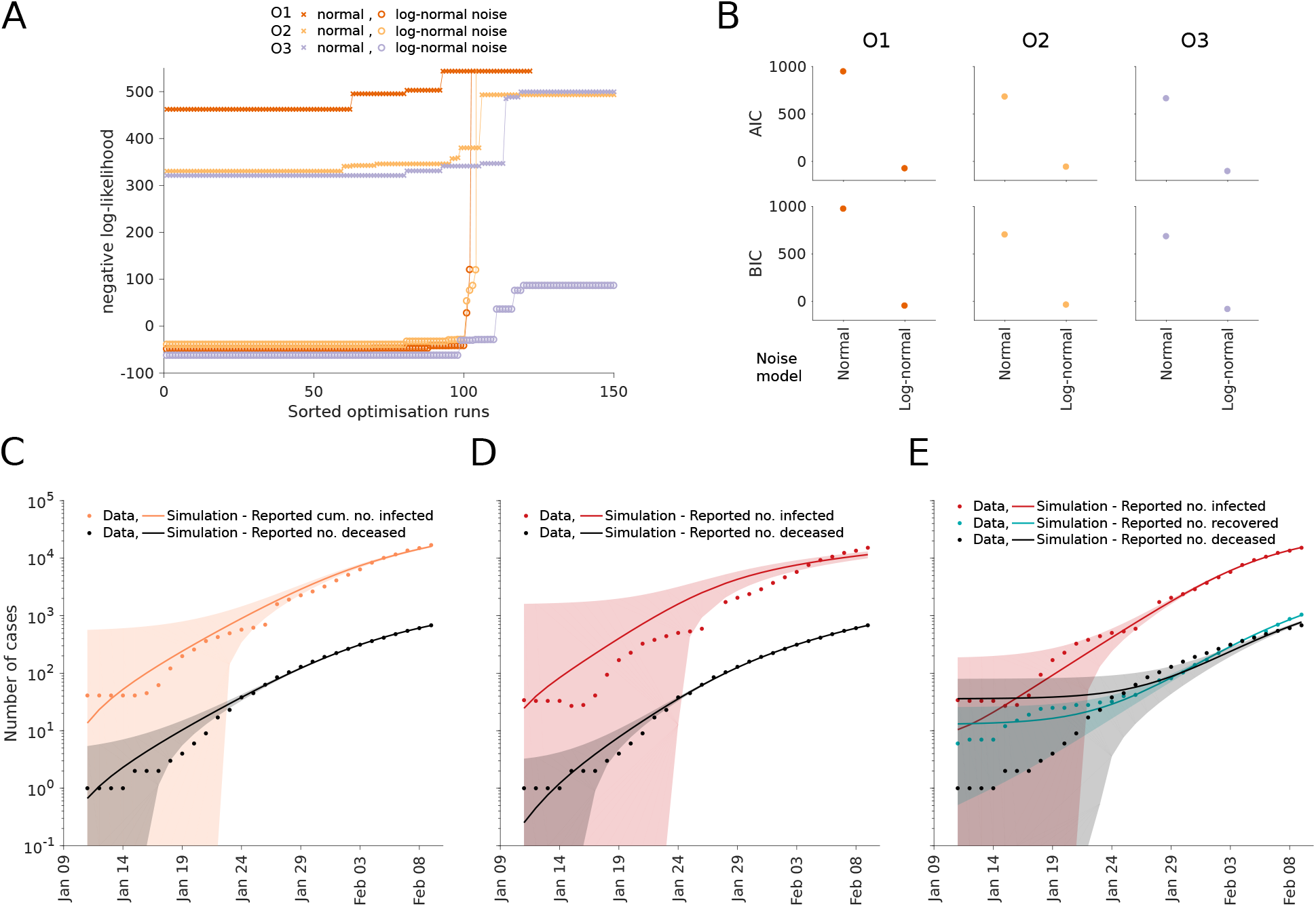
Optimisation and model selection results for observable and noise model comparison. (A) Waterfall plots for the best 150 out of 200 multi-start runs. The optimisation runs are sorted by increasing negative log-likelihood value. (B) AIC and BIC values. (C,D,E) Fitting results for observation scenarios O1, O2, and O3 assuming normally distributed noise. The simulation for the maximum likelihood estimate (line) and interval for +*/−* one standard deviation of the inferred noise level (shaded area) is depicted.

**Figure S2:**
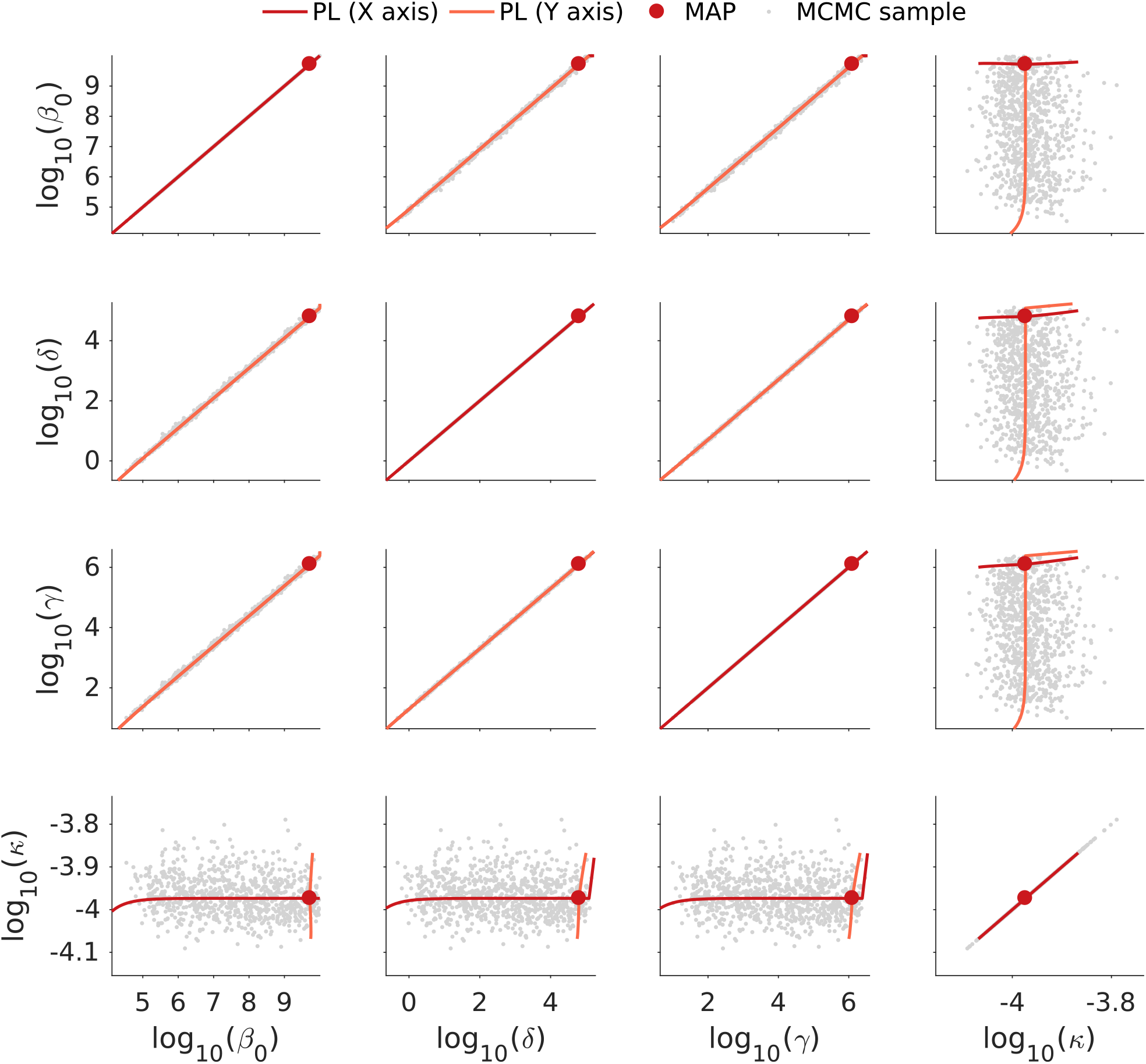
Parameter correlation for model O1. The scatter plot matrix for the MCMC samples are depicted. The maximum a posterior estimate (MAP) and the profile likelihoods with respect to the parameter in the x-axis and y-axis are indicated.

**Figure S3:**
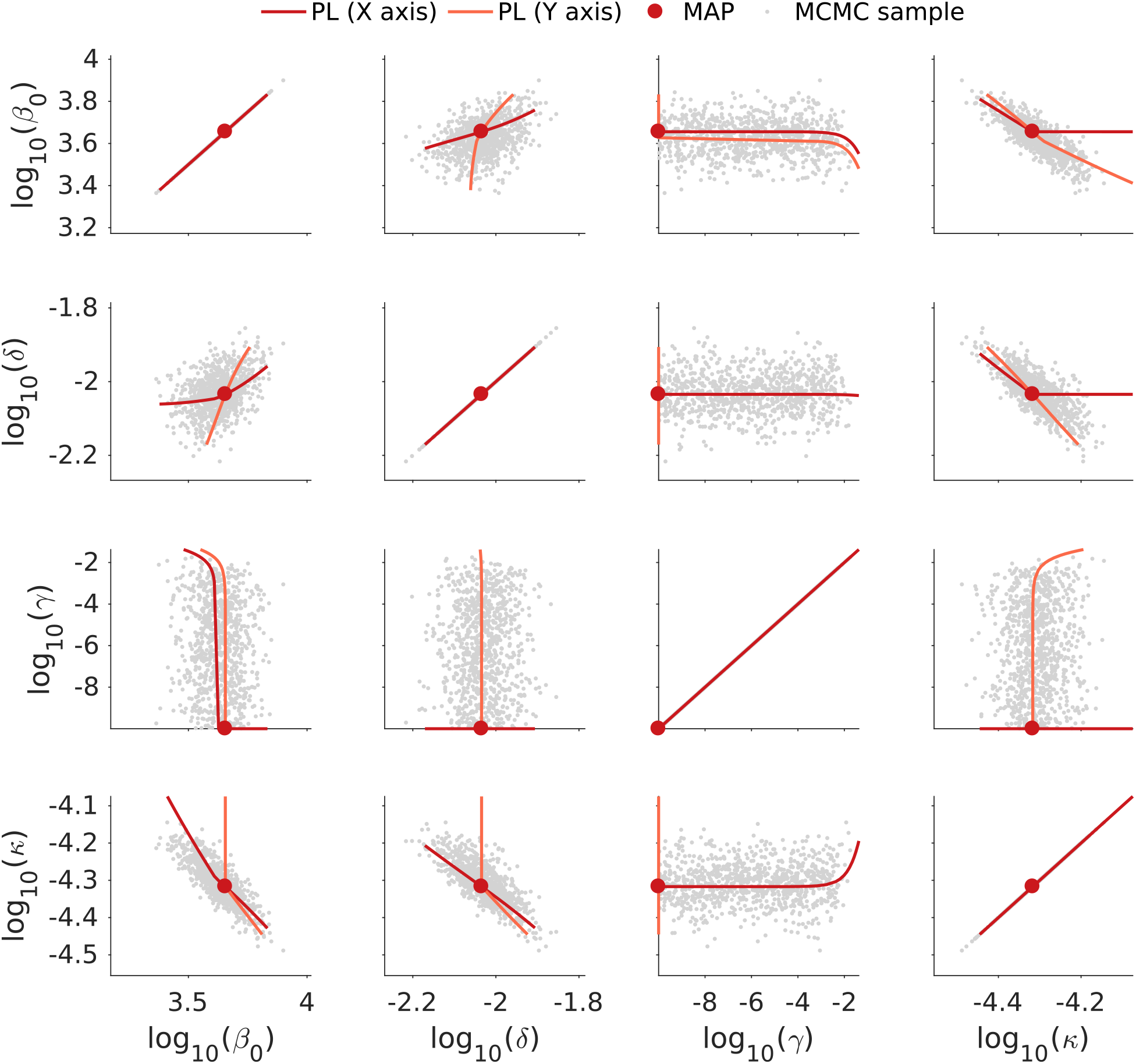
Parameter correlation for model O2. The scatter plot matrix for the MCMC samples are depicted. The maximum a posterior estimate (MAP) and the profile likelihoods with respect to the parameter in the x-axis and y-axis are indicated.

**Figure S4:**
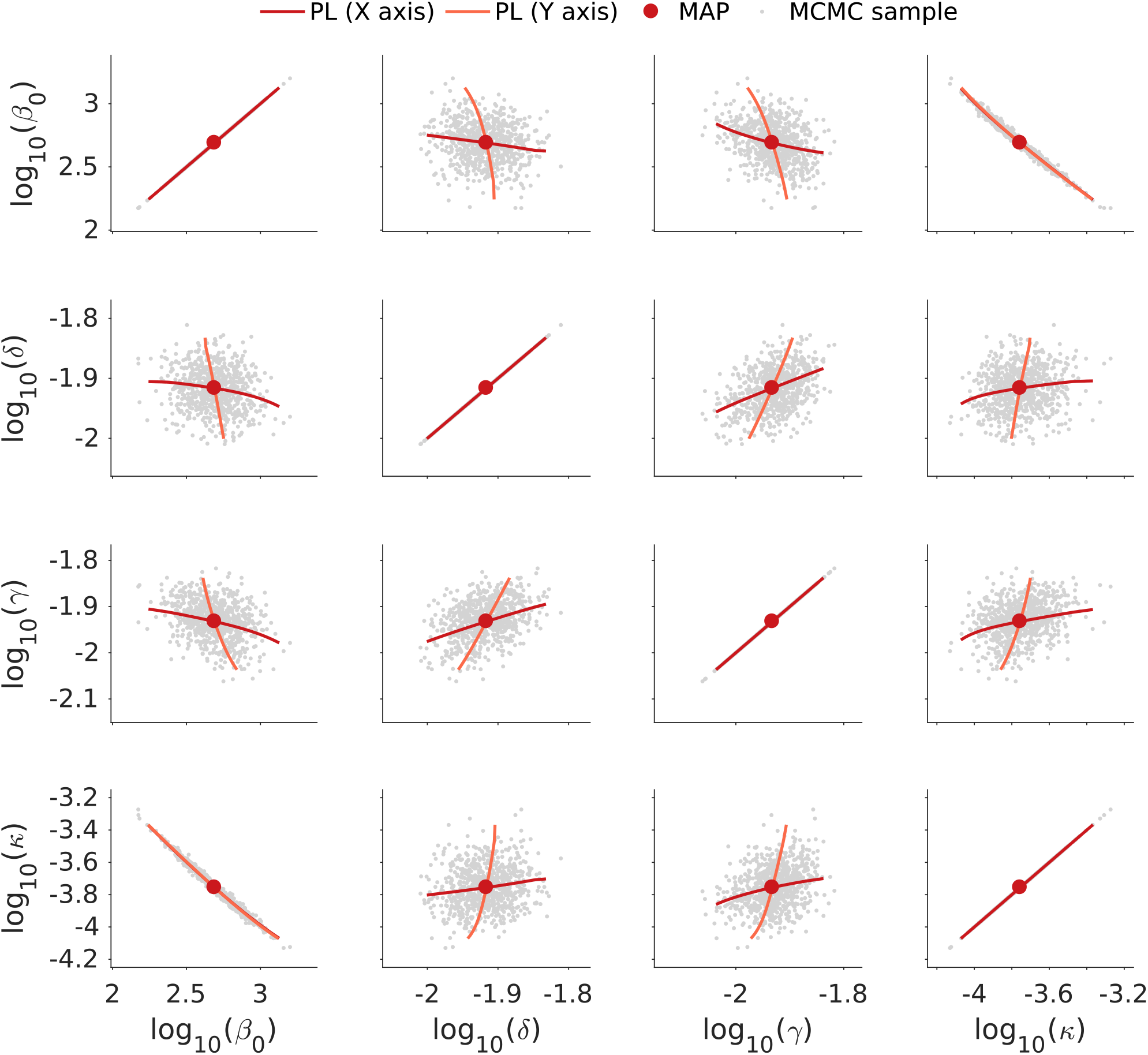
Parameter correlation for model O3. The scatter plot matrix for the MCMC samples are depicted. The maximum a posterior estimate (MAP) and the profile likelihoods with respect to the parameter in the x-axis and y-axis are indicated.

## References

[1] Yang, W., Cowling, B. J., Lau, E. H. Y. & Shaman, J. Forecasting Influenza Epidemics in Hong Kong. PLoS Comput Biol 11, e1004383 (2015).

[2] Reich, N. G. et al.. Challenges in Real-Time Prediction of Infectious Disease: A Case Study of Dengue in Thailand. PLoS Neglect Trop D 10, e0004761 (2016).

[3] Shaman, J., Yang, W. & Kandula, S. Inference and Forecast of the Current West African Ebola Outbreak in Guinea, Sierra Leone and Liberia. PLoS Curr 6 (2014).

[4] Chowell, G. et al.. Using Phenomenological Models to Characterize Transmissibility and Forecast Patterns and Final Burden of Zika Epidemics. PLoS Curr 8 (2016).

[5] Ferguson, N. et al.. Report 9: Impact of non-pharmaceutical interventions (NPIs) to reduce COVID19 mortality and healthcare demand (2020). URL https://www.imperial.ac.uk/media/imperial-college/medicine/sph/ide/gida-fellowships/Imperial-College-COVID19-NPI-modelling-16-03-2020.pdf.

[6] Boldog, P. et al.. Risk Assessment of Novel Coronavirus COVID-19 Outbreaks Outside China. J Clin Med 9, 571 (2020).

[7] Doms, C., Kramer, S. C. & Shaman, J. Assessing the Use of Influenza Forecasts and Epidemiological Modeling in Public Health Decision Making in the United States. Sci Rep 8, 1–7 (2018).

[8] Kermack, W. O., McKendrick, A. G. & Walker, G. T. A Contribution to the Mathematical Theory of Epidemics. P Roy Soc A-Math Phy 115, 700–721 (1927).

[9] Hethcote, H. W. The Mathematics of Infectious Diseases. SIAM Rev 42 (2000).

[10] Brauer, F. & Castillo-Chavez, C. Epidemic Models. In Brauer, F. & Castillo-Chavez, C. (eds.) Mathematical Models in Population Biology and Epidemiology, Texts in Applied Mathematics, 345–409 (Springer, New York, NY, 2012).

[11] Chalub, F. A. C. C. & Souza, M. O. The SIR epidemic model from a PDE point of view. Math Comput Model 53, 1568–1574 (2011).

[12] Lotfi, E. M., Maziane, M., Hattaf, K. & Yousfi, N. Partial Differential Equations of an Epidemic Model with Spatial Diffusion. Int J Partial Differ Equ 2014 (2014).

[13] Dargatz, C., Georgescu, V. & Held, L. Stochastic Modelling of the Spatial Spread of Influenza in Germany. Austrian J Stat 35, 5–20 (2006).

[14] Greenwood, P. E. & Gordillo, L. F. Stochastic Epidemic Modeling. In Chowell, G., Hyman, J. M., Bettencourt, L. M. A. & Castillo-Chavez, C. (eds.) Mathematical and Statistical Estimation Approaches in Epidemiology, 31–52 (Springer Netherlands, Dordrecht, 2009).

[15] Allen, L. J. S. An Introduction to Stochastic Processes with Applications to Biology (Chapman and Hall/CRC, 2010).

[16] Britton, T. Stochastic epidemic models: A survey. Math Biosci 225, 24–35 (2010).

[17] Isham, V. Stochastic Models for Epidemics (Oxford University Press, 2007).

[18] Epstein, J. M. & Axtell, R. Growing Artificial Societies: Social Science from the Bottom Up. MIT Press Books (1996). ISBN: 9780262550253.

[19] Bruch, E. & Atwell, J. Agent-Based Models in Empirical Social Research. Sociol Methods Res 44, 186–221 (2015).

[20] Birrell, P. J. et al. Bayesian modeling to unmask and predict influenza A/H1N1pdm dynamics in London. PNAS 108, 18238–18243 (2011).

[21] Brookhart, M. A., Hubbard, A. E., van der Laan, M. J., Colford, J. M. & Eisenberg, J.N. S. Statistical Estimation of Parameters in a Disease Transmission Model: Analysis of a Cryptosporidium Outbreak. Stat Med 21, 3627–3638 (2002).

[22] Weidemann, F., Dehnert, M., Koch, J., Wichmann, O. & Höhle, M. Bayesian parameter inference for dynamic infectious disease modelling: Rotavirus in Germany. Stat Med 33, 1580–1599 (2014).

[23] Chatzilena, A., van Leeuwen, E., Ratmann, O., Baguelin, M. & Demiris, N. Contemporary Statistical Inference for Infectious Disease Models Using Stan. Epidemics 29, 100367 (2019).

[24] Farah, M., Birrell, P., Conti, S. & Angelis, D. D. Bayesian Emulation and Calibration of a Dynamic Epidemic Model for A/H1N1 Influenza. J Am Stat Assoc 109, 1398–1411 (2014).

[25] Akaike, H. Information theory and an extension of the maximum likelihood principle. In 2nd International Symposium on Information Theory, Tsahkadsor, Armenian SSR, vol. 1, 267–281 (Akademiai Kiado, 1973).

[26] Schwarz, G. Estimating the dimension of a model. Ann Stat 6, 461–464 (1978).

[27] Kass, R. E. & Raftery, A. E. Bayes factors. J Am Stat Assoc 90, 773–795 (1995).

[28] Gralinski, L. E. & Menachery, V. D. Return of the coronavirus: 2019-nCoV. Viruses 12, 135 (2020).

[29] World Health Organization. Novel Coronavirus (2019-nCoV) situation report 7 (2020). URL https://www.who.int/docs/default-source/coronaviruse/situation-reports/20200127-sitrep-7-2019--ncov.pdf.

[30] Tedros, A. G. WHO Director-General’s opening remarks at the media briefing on COVID-19 - 11 March 2020 (2020). URL https://www.who.int/dg/speeches/detail/who-director-general-s-opening-remarks-at-the-media-briefing-on-covid-1911-march-2020.

[31] Shao, P. & Shan, Y. Beware of asymptomatic transmission: Study on 2019-nCoV prevention and control measures based on extended SEIR model. bioRxiv (2020). doi: 10.1101/2020.01.28.923169.

[32] Chen, T.-M. et al. A mathematical model for simulating the phase-based transmissibility of a novel coronavirus. Infect Dis Poverty 9, 24 (2020).

[33] Li, R. et al. Substantial undocumented infection facilitates the rapid dissemination of novel coronavirus (SARS-CoV2). Science (2020).

[34] Ming, W.-K., Huang, J. & Zhang, C. J. P. Breaking down of the healthcare system: Mathematical modelling for controlling the novel coronavirus (2019-nCoV) outbreak in Wuhan, China. bioRxiv (2020). doi:10.1101/2020.01.27.922443.

[35] Read, J. M., Bridgen, J. R., Cummings, D. A., Ho, A. & Jewell, C. P. Novel coronavirus 2019-nCoV: Early estimation of epidemiological parameters and epidemic predictions. medRxiv (2020). doi:10.1101/2020.01.23.20018549.

[36] Koo, J. R. et al. Interventions to mitigate early spread of SARS-CoV-2 in Singapore: A modelling study. Lancet Infect Dis (2020).

[37] Neher, R. A., Dyrdak, R., Druelle, V., Hodcroft, E. B. & Albert, J. Potential impact of seasonal forcing on a SARS-CoV-2 pandemic. Swiss Med Wkly 150 (2020).

[38] Li, Q. et al. Early Transmission Dynamics in Wuhan, China, of Novel Coronavirus–Infected Pneumonia. New Engl J Med 382, 1199–1207 (2020).

[39] Zhao, S. et al. Preliminary Estimation of the Basic Reproduction Number of Novel Coronavirus (2019-nCoV) in China, from 2019 to 2020: A Data-Driven Analysis in the Early Phase of the Outbreak. Int J Infect Dis 92, 214–217 (2020).

[40] Tian, H. et al. An investigation of transmission control measures during the first 50 days of the COVID-19 epidemic in China. Science (2020).

[41] Liu, Z., Magal, P., Seydi, O. & Webb, G. Predicting the cumulative number of cases for the COVID-19 epidemic in China from early data. medRxiv (2020). doi:10.1101/2020.03.11.20034314.

[42] Nordt, C. & Herdener, M. A pragmatic model to forecast the COVID-19 epidemic in different countries and allowing for daily updates. medRxiv (2020). doi:10.1101/2020.04.07.20056481.

[43] Jenny, P., Jenny, D. F., Gorji, H., Arnoldini, M. & Hardt, W.-D. Dynamic Modeling to Identify Mitigation Strategies for Covid-19 Pandemic. medRxiv (2020). doi:10.1101/2020.03.27.20045237.

[44] Health Commission of Hubei Province (2020). URL http://wjw.hubei.gov.cn/.

[45] Wuhan Municipal Health Commission (2020). URL http://wjw.wuhan.gov.cn/.

[46] Zhao, S. et al. Estimating the unreported number of novel coronavirus (2019-nCoV) cases in China in the first half of January 2020: a data-driven modelling analysis of the early outbreak. J Clin Med 9, 388 (2020).

[47] Wang, H. et al. Phase-adjusted estimation of the number of coronavirus disease 2019 cases in Wuhan, China. Cell Discov 6, 1–8 (2020).

[48] Roosa, K. et al. Real-time forecasts of the COVID-19 epidemic in China from February 5th to February 24th, 2020. Infect Dis Model 5, 256–263 (2020).

[49] Peng, L., Yang, W., Zhang, D., Zhuge, C. & Hong, L. Epidemic analysis of COVID-19 in China by dynamical modeling. arXiv preprint 2002.06563 (2020).

[50] Chinese Center for Disease Control and Prevention (2020). URL http://www.chinacdc.cn/en/.

[51] Capasso, V. Mathematical Structures of Epidemic Systems. Lecture Notes in Biomathematics (Springer-Verlag, 1993).

[52] Chowell, G. Fitting dynamic models to epidemic outbreaks with quantified uncertainty: a primer for parameter uncertainty, identifiability, and forecasts. Infect Dis Model 2, 379–398 (2017).

[53] World Health Organization. Novel Coronavirus (2019-nCoV) situation report 30 (2020). URL https://www.who.int/docs/default-source/coronaviruse/situation-reports/20200219-sitrep-30-covid-19.pdf?sfvrsn=3346b04f2.

[54] Backer, J. A., Klinkenberg, D. & Wallinga, J. Incubation period of 2019 novel coronavirus (2019-nCoV) infections among travellers from Wuhan, China, 20–28 January 2020. Eurosurveillance 25, 2000062 (2020).

[55] European Centre for Disease Prevention and Control. Outbreak of novel coronavirus disease 2019 (COVID-19): increased transmission globally – fifth update (2020). URL https://www.ecdc.europa.eu/sites/default/files/documents/RRA-outbreak-novel-coronavirus-disease-2019-increase-transmission-globally-COVID-19.pdf.

[56] Amanat, F. & Krammer, F. SARS-CoV-2 Vaccines: Status Report. Immunity (2020). doi:10.1016/j.immuni.2020.03.007.

[57] Maier, B. F. & Brockmann, D. Effective containment explains subexponential growth in recent confirmed COVID-19 cases in China. Science (2020). doi:10.1126/science.abb4557.

[58] Barbarossa, M. V. et al. A first study on the impact of current and future control measures on the spread of COVID-19 in Germany. medRxiv (2020). doi:10.1101/2020.04.08.20056630.

[59] Salim, N. et al. Covid-19 epidemic in Malaysia: Impact of lock-down on infection dynamics. medRxiv (2020). doi:10.1101/2020.04.08.20057463.

[60] Berk, S. H. & Kadyrov, S. Purely data-driven exploration of COVID-19 pandemic after three months of the outbreak. medRxiv (2020). doi:10.1101/2020.04.08.20057638.

[61] He, S., Tang, S. & Rong, L. A discrete stochastic model of the COVID-19 outbreak: Forecast and control. Math Biosci Eng 17, 2792–2804 (2020).

[62] Halloran, M. E. et al. Modeling targeted layered containment of an influenza pandemic in the United States. P Natl Acad Sci 105, 4639–4644 (2008).

[63] Wu, J. T. et al. Estimating clinical severity of COVID-19 from the transmission dynamics in Wuhan, China. Nat Med 26, 506–510 (2020).

[64] Nishiura, H. et al. Estimation of the asymptomatic ratio of novel coronavirus infections (COVID-19). Int J Infect Dis (2020).

[65] Khailaie, S. et al. Estimate of the development of the epidemic reproduction number Rt from Coronavirus SARS-CoV-2 case data and implications for political measures based on prognostics. medRxiv (2020). doi:10.1101/2020.04.04.20053637.

[66] Lauer, S. A. et al. The incubation period of coronavirus disease 2019 (COVID-19) from publicly reported confirmed cases: estimation and application. Ann Intern Med (2020).

[67] Zhou, F. et al. Clinical course and risk factors for mortality of adult inpatients with COVID-19 in Wuhan, China: a retrospective cohort study. Lancet 395, 1054–1062 (2020).

[68] Wölfel, R. et al. Virological assessment of hospitalized patients with COVID-2019. Nature 1–10 (2020). doi:10.1038/s41586-020-2196-x.

[69] Bayerisches Staatsministerium für Wissenschaft und Kunst. Pressemitteilung Nr. 072: Gemeinsam gegen COVID-19: Münchner Tropeninstitut beginnt Stichprobenanalyse zur Verbreitung der Corona-Pandemie und zur Wirksamkeit von Gegenmaßnahmen. URL https://www.stmwk.bayern.de/pressemitteilung/11894/gemeinsam-gegen-covid-19-muenchner-tropeninstitut-beginnt-stichprobenanalyse-zur-verbreitung-der-corona-pandemie-und-zur-wirksamkeit-von-gegenmassnahmen.html. Accessed: 2020-04-14.

[70] Business Insider, Ashley Collman (2020). URL https://www.businessinsider.com/5-million-left-wuhan-before-coronavirus-quarantine-2020-1?r=DE&IR=T.

[71] European Centre for Disease Prevention and Control. Event background COVID-19 (2020). URL https://www.ecdc.europa.eu/en/novel-coronavirus/event-background-2019.

[72] Raue, A. et al. Structural and practical identifiability analysis of partially observed dynamical models by exploiting the profile likelihood. Bioinformatics 25, 1923–1929 (2009).

[73] Kreutz, C., Raue, A., Kaschek, D. & Timmer, J. Profile likelihood in systems biology. FEBS J 280, 2564–2571 (2013).

[74] Raue, A. et al. Data2Dynamics: a modeling environment tailored to parameter estimation in dynamical systems. Bioinformatics 31, 3558–3560 (2015).

[75] Hass, H., Kreutz, C., Timmer, J. & Kaschek, D. Fast integration-based prediction bands for ordinary differential equation models. Bioinformatics 32, 1204–1210 (2016).

[76] Meeker, W. Q. & Escobar, L. A. Teaching about approximate confidence regions based on maximum likelihood estimation. Am Stat 49, 48–53 (1995).

[77] Ballnus, B. et al. Comprehensive benchmarking of Markov chain Monte Carlo methods for dynamical systems. BMC Syst Biol 11, 63 (2017).

[78] Stapor, P. et al. PESTO: Parameter EStimation TOolbox. Bioinformatics 34, 705–707 (2018).

[79] Geweke, J. Evaluating the accuracy of sampling-based approaches to the calculation of posterior moments. In Bernardo, J. M., Smith, A. F. M., Dawid, A. P. & Berger, J. O. (eds.) Bayesian Statistics, vol. 4, 169–193 (University Press Oxford, 1992).

[80] Burnham, K. P. & Anderson, D. R. Model selection and multimodel inference: A practical information-theoretic approach (Springer, New York, NY, 2002), 2nd edn.

[81] Fröhlich, F., Kaltenbacher, B., Theis, F. J. & Hasenauer, J. Scalable parameter estimation for genome-scale biochemical reaction networks. PLoS Comput Biol 13, e1005331 (2017).

[82] Fröhlich, F., Theis, F. J., Rädler, J. O. & Hasenauer, J. Parameter estimation for dynamical systems with discrete events and logical operations. Bioinformatics 33, 1049–1056 (2017).

[83] Serban, R. & Hindmarsh, A. C. CVODES: The sensitivity-enabled ODE solver in SUN-DIALS. In ASME 2005 International Design Engineering Technical Conferences and Computers and Information in Engineering Conference, vol. 6, 257–269 (ASME, 2005).

[84] Schmiester, L. et al. PEtab – interoperable specification of parameter estimation problems in systems biology. arXiv preprint 2004.01154 (2020).

[85] Hucka, M. et al. The systems biology markup language (SBML): A medium for representation and exchange of biochemical network models. Bioinformatics 19, 524–531 (2003).

